# Associations between lack of social support and food insecurity: A cross-sectional analysis of the 2024 BRFSS

**DOI:** 10.64898/2026.05.24.26353990

**Authors:** Eashwar Krishna, Neha Shanavas, Fatima Mir, Ahan Kothapeta, Carlos Duluc, Roma Kale, Priya Bheemanakunta, Eshita Mathur

## Abstract

**Introduction:** Growing evidence shows a strong link between social connection and food security, with social support often serving as a buffer against food hardship and its mental health impacts. However, most research on this relationship comes from international studies. The few studies conducted in the United States rely on outdated data, focus strictly on older adults, or are limited to specific geographic regions. To address these limitations and provide a clearer national picture, this study uses recent, individual-level data from a diverse U.S. sample to examine how social isolation and food insecurity interact.

**Materials & Methods:** Cross-sectional secondary data analysis conducted on Behavioral Risk Factor Surveillance System (BRFSS) data from 2024, collected via a nationwide telephone survey. The sample consisted of Adults (n = 190,577) aged 18-80 years old (72.3% non-Hispanic White). Food insecurity was defined as responding always, usually, or sometimes to “During the past 12 months how often did the food that you bought not last, and you didn’t have money to buy more?” Social support was measured using a BRFSS item assessing the frequency with which respondents received the social and emotional support they needed. Adjusted logistic regression models were used to assess the relationship between these variables while controlling for a wide variety of demographic, socioeconomic, and health status factors.

**Results:** Individuals who reported only “sometimes” receiving the social and emotional support they need were more likely to report food insecurity as compared to those who “always” receive such support (aOR = 1.75; 95% CI 1.56, 1.96).

**Discussion:** These findings indicate that decreased social support may put individuals at higher risk of food insecurity. Future work should seek to understand the mechanisms of this association to inform targeted policy and other interventional programs.

## Introduction

Social isolation has been increasingly identified as a key mechanistic pathway linking food insecurity to adverse mental health outcomes. Emerging evidence suggests that individuals experiencing food insecurity are more likely to report higher levels of social isolation, which in turn contributes to elevated depressive symptoms. In a cross-sectional study of rural Midwestern veterans, social isolation was found to significantly mediate the association between both short-term (30-day) and long-term (12-month) food insecurity and depressive symptoms, indicating that part of the mental health burden of food insecurity operates through disrupted or diminished social connectedness.^1^ Notably, this mediating effect was observed across time frames of food insecurity exposure, suggesting that both acute and chronic deprivation may exert psychological effects through similar social pathways. These findings are consistent with broader stress-process models, which propose that structural stressors such as food insecurity reduce access to supportive social networks, thereby increasing vulnerability to psychological distress. Together, this evidence positions social isolation not merely as a co-occurring correlate of food insecurity, but as a potential intermediary mechanism through which food-related hardship translates into worse mental health outcomes.

A growing body of research internationally has demonstrated a relationship between social support and food insecurity, suggesting that social and emotional resources may play a role against food insecurity. Studies conducted in diverse global contexts provide insight into how different forms of support can influence food security. Higher levels of social support have been linked with significantly lower odds of moderate and severe household food insecurity, with effect estimates indicating a consistent positive relationship across severity levels of food insecurity.^2^ In this study of households in Rio de Janeiro, individuals reporting stronger social support were less likely to report food insecurity. Additionally, evidence suggests that this relationship operates across various forms of support, including affective, material, informational and social support, all of which has been linked to reduced chances of food insecurity.^3^ This framework emphasizes that not only practical forms, but emotional forms of support can also contribute to improved food security.

More research in medically vulnerable populations suggests that higher levels of informational, social interaction and support are inversely related with mild and severe food insecurity.^4^ In families of children with sickle cell disease, access to informational guidance and social engagement was linked to lower odds of food insecurity, highlighting how various forms of social support may help alleviate financial and caregiving burdens. Across these studies, the consistency of the relationship between social support and food security supports the importance of social connections in shaping food access and stability.

Despite the strength of these findings, a key limitation for these studies is that they are primarily derived from international populations, outside of the United States. Differences in social infrastructure, healthcare, and economic safety nets may influence both availability and effectiveness of social support. This may limit the generalizability of these results, so it remains necessary to evaluate whether this relationship exists within the U.S. context.

Few studies exist evaluating the relationship between social connection and food insecurity within the United States. One national study assessed Americans over 65 years of age and found that elderly adults with more social strain and less emotional support were at higher risk of food insecurity.^5^ Though this finding suggests a critical link, the study’s scope was limited to a specific elderly age cohort and relied on data from 2012 and 2013 with a smaller sample of 1164 Americans. Given the shifting economic and social landscape in the last decade, these results may not be generalizable to the current American public. Therefore, there remains a pressing need for contemporary research that examines this relationship across a diverse age range at the national level.

Recent research has furthered this inquiry at a regional level. Krishna et al. demonstrated that among Connecticut residents, self-reported social and emotional support was significantly associated with reduced food insecurity and with greater access to affordable fresh produce.^6^ However, a major limitation of this study was that data was sourced at the town-level and only focused on Connecticut residents. This methodology could leave the findings susceptible to local geographical confounders. To address these limitations and increase generalizability, the present study uses individual-level data from a national American sample to confirm the association between social isolation and food insecurity.

## Methods

This study used data from the 2024 Behavioral Risk Factor Surveillance System (BRFSS), a nationally representative, cross-sectional survey conducted by the Centers for Disease Control and Prevention (CDC) that collects information on health-related risk behaviors, chronic conditions, and use of preventive services among U.S. adults.^7^ The analytic sample included 190,577 respondents with complete data on food insecurity, social support, and relevant covariates.

Food insecurity was assessed using a validated single-item BRFSS measure, coded as a binary outcome: food secure versus food insecure. Respondents who responded with always, usually and sometimes to the question “During the past 12 months how often did the food that you bought not last, and you didn’t have money to buy more?” were classified as food insecure. Social support was measured using a BRFSS item assessing the frequency with which respondents received the social and emotional support they needed. Responses were coded into a four-level ordinal variable: always, usually, sometimes, and rarely/never. For bivariate analyses, inadequate social support was defined as responding sometimes, rarely, or never.

Demographic covariates included age, sex, race/ethnicity (White Non-Hispanic, Black Non-Hispanic, Hispanic, Other Non-Hispanic, Multiracial Non-Hispanic), and marital status. Socioeconomic variables included education, employment status, annual household income, and Supplemental Nutrition Assistance Program (SNAP) receipt. Health-related variables included self-rated general health, number of physically and mentally unhealthy days in the past 30 days, and a history of depression. Geographic location was categorized as urban or rural.

Weighted proportions and means were calculated to describe the distribution of food insecurity and social support overall and stratified by race/ethnicity. Bivariate associations between social support and food insecurity were tested using the Rao-Scott chi-square test, accounting for the BRFSS complex survey design.

Three hierarchical logistic regression models were estimated to assess the association between social support and food insecurity:

- Model 1: Unadjusted association between social support and food insecurity.
- Model 2: Adjusted for demographic characteristics (sex, age, race/ethnicity, and marital status).
- Model 3: Fully adjusted model, including demographics, socioeconomic status (education, employment, income, SNAP), health status (general health, physical and mental unhealthy days, depression), and geography.

Odds ratios (ORs) and adjusted odds ratios (aORs) with 95% confidence intervals (CIs) were reported, with always receiving social support as the reference category.

To explore potential effect modification by race/ethnicity, stratified analyses were conducted, estimating aORs for food insecurity at each level of social support within racial/ethnic groups. Potential effect modification by binary rural/urban status (based on the 6-tier NCHS classification) was explored in this manner. Additionally, ordinal logistic regression models were used as sensitivity analyses to examine whether social support predicted severity of food insecurity, with similar covariate adjustment.

All analyses incorporated BRFSS sampling weights, strata, and primary sampling units to account for the complex survey design and ensure nationally representative estimates. Analyses were conducted in R (version 4.5.1). Statistical significance was defined as p < 0.05 for all tests.

## Results

The two primary analysis variables, social support (four-stage ordinal) and food insecurity (binary), were compared between the racial categories provided in the survey and also to each other. A clear trend was observed between these two variables. Among those who reported always receiving the social support they needed, only 10.5% reported being food insecure. In stark contrast, among those who rarely/never received social support, 30% of the sample reported food insecurity, and these trends demonstrated significance via a Rao-Scott bivariate test (X² = 7,588, p < .001), although this is to be expected in a sample of this size (n=190,577). Significant differences from the average food insecurity rate were observed for White, non-Hispanic Black, and Hispanic respondents; the difference for multiracial respondents approached significance but did not reach it. Social support differences were observed across all racial categories. The sample consisted of 72.3% non-Hispanic White, 8.4% non-Hispanic Black, 10.2% Hispanic, 5.2% other non-Hispanic, and 2.3% non-Hispanic multiracial.

Three logistic regression models were estimated to assess the association between social/emotional support and food insecurity. Model 1 was unadjusted, Model 2 controlled for demographic characteristics (sex, age, race/ethnicity, and marital status), and Model 3 additionally controlled for socioeconomic status (education, employment, income, and SNAP receipt), health status (general health, physical and mental unhealthy days, and depression), and geography (urban/rural status). Odds ratios for the primary exposure across all three models are presented in Table 2, and full Model 3 results are presented in Table 3.

**Table 1:** Food Insecurity and Inadequate Social Support by Race/Ethnicity.

| Race/Ethnicity | Food Insecure (95% CI) | Inadequate Support (95% CI) |
| --- | --- | --- |
| White, Non-Hispanic | 9.2% (8.9-9.5) | 20.0% (19.6-20.0) |
| Black, Non-Hispanic | 22.1% (21.1-23.0) | 31.0% (29.8-32.0) |
| Hispanic | 25.5% (24.3-26.0) | 27.4% (26.0-28.7) |
| Other, Non-Hispanic | 13.5% (11.8-15.0) | 30.6% (28.1-33.0) |
| Multiracial, Non-Hispanic | 17.1% (14.8-19.0) | 29.7% (26.8-32.0) |
| Overall Population | 14.6% (14.3-15.0) | 24.1% (23.7-24.0) |
| Rao-Scott X<sup>2</sup> | 7247.9*** | 2281.6*** |

\*\*\* $p < .001$ . “Inadequate” social support defined as respondents answering that they sometimes, rarely, or never got the emotional and/or social support they needed. Significant Rao-Scott values indicate differences between groups beyond chance.

**Table 2.**
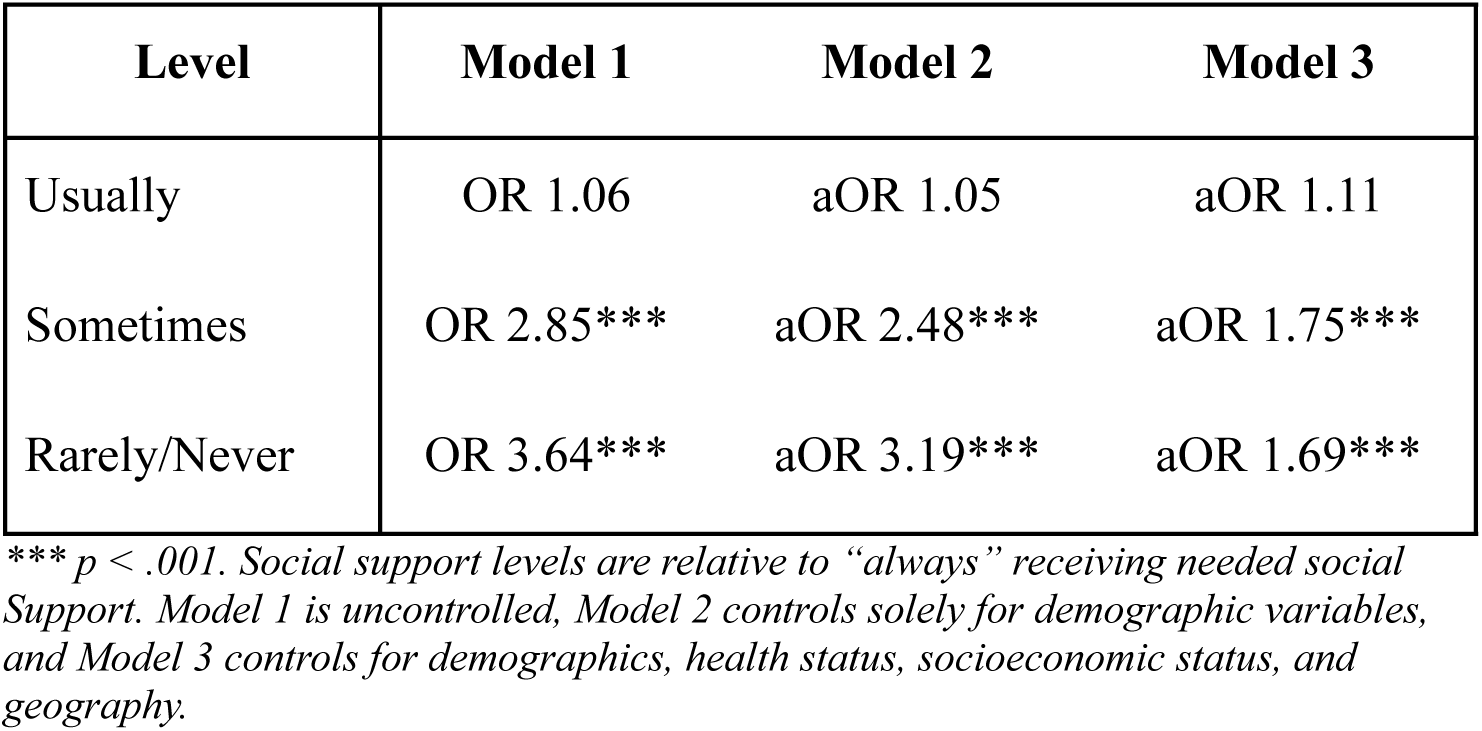
Adjusted Odds Ratios for Food Insecurity.

| Level | Model 1 | Model 2 | Model 3 |
| --- | --- | --- | --- |
| Usually | OR 1.06 | aOR 1.05 | aOR 1.11 |
| Sometimes | OR 2.85*** | aOR 2.48*** | aOR 1.75*** |
| Rarely/Never | OR 3.64*** | aOR 3.19*** | aOR 1.69*** |

\*\*\* $p < .001$ . Social support levels are relative to “always” receiving needed social support. Model 1 is uncontrolled, Model 2 controls solely for demographic variables, and Model 3 controls for demographics, health status, socioeconomic status, and geography.

**Table 3.** Fully-adjusted multivariable logistic regression results (Model 3)

| Variable Category | Term | aOR (95% CI) |
| --- | --- | --- |
| **Social Support** | Usually | 1.11 (1.00–1.24) |
|  | Sometimes | 1.75 (1.56–1.96)*** |
|  | Rarely/Never | 1.69 (1.47–1.95)*** |
| **Sex** | Female | 1.23 (1.13–1.34)*** |
| **Age Group** | 25–34 | 1.05 (0.89–1.25) |
|  | 35–44 | 1.04 (0.87–1.24) |
|  | 45–54 | 0.86 (0.71–1.04) |
|  | 55–64 | 0.68 (0.56–0.83)*** |
|  | 65+ | 0.47 (0.38–0.59)*** |
| **Race/Ethnicity** | Black, Non-Hispanic | 1.89 (1.71–2.10)*** |
|  | Hispanic | 1.57 (1.39–1.76)*** |
|  | Other, Non-Hispanic | 1.51 (1.23–1.87)*** |
|  | Multiracial, Non-Hispanic | 1.16 (0.91–1.47) |
| **Marital Status** | Never Married/Unmarried | 0.82 (0.73–0.92)*** |
|  | Divorced | 0.89 (0.79–1.01) |
|  | Separated | 1.22 (0.98–1.53) |
|  | Widowed | 0.91 (0.78–1.06) |
| **Education** | HS Graduate | 0.70 (0.62–0.81)*** |
|  | Some College | 0.59 (0.51–0.68)*** |
|  | College Graduate | 0.45 (0.38–0.52)*** |
| **Employment** | Unemployed | 1.39 (1.19–1.63)*** |
|  | Retired | 0.80 (0.68–0.94)** |
|  | Unable to Work | 1.21 (1.04–1.40)* |
|  | Homemaker | 0.67 (0.54–0.84)*** |
|  | Student | 0.70 (0.54–0.91)** |
| **Annual Income** | \$15,000–\$24,999 | 0.96 (0.83–1.12) |
| | \$25,000–\$34,999 | 0.65 (0.56–0.77)*** |
| | \$35,000–\$49,999 | 0.43 (0.37–0.51)*** |
| | \$50,000–\$74,999 | 0.24 (0.20–0.28)*** |
| | \$75,000+ | 0.14 (0.11–0.17)*** |
| **SNAP Benefits** | Yes | 2.22 (1.99–2.48)*** |
| **Health Factors** | Fair/Poor Gen. Health | 1.45 (1.30–1.62)*** |
|  | Physical Health Days | 1.01 (1.01–1.02)*** |
|  | Mental Health Days | 1.02 (1.02–1.03)*** |
|  | Depression (Yes) | 1.11 (1.00–1.23)* |
| **Geography** | Rural | 0.96 (0.86–1.07) |

Reference Categories: Social Support (Always), Sex (Male), Age (18–24), Race (White Non-Hispanic), Marital Status (Married), Education (<HS), Employment (Employed), Income (<\$15k), SNAP (No), General Health (Good/Very Good/Excellent), Rurality (Urban), Depression (No). \*\*\* $p < .001$ , \*\* $p < .01$ , \* $p < 0.5$

In the unadjusted model, receiving social support sometimes (OR 2.85, p < .001) or rarely/never (OR 3.64, p < .001) was associated with substantially elevated odds of food insecurity relative to always receiving support. The usually versus always contrast was not significant in any model (Model 3: aOR 1.11, p = .058). After adjustment for demographic covariates in Model 2, the sometimes and rarely/never estimates attenuated modestly (aOR 2.48 and 3.19, respectively), and attenuated further in the fully adjusted Model 3 (aOR 1.75 and 1.69, respectively), indicating that a substantial portion of the crude association is explained by socioeconomic and health-related factors. Nevertheless, both estimates remained large and statistically significant (p < .001), supporting an independent association between inadequate social support and food insecurity net of all covariates.

Among the covariates included in Model 3, income was the strongest predictor of food insecurity, with each successive income bracket associated with progressively lower odds; those earning $75,000 or more had 86% lower odds of food insecurity than those earning less than $15,000 (aOR 0.14, 95% CI 0.11-0.17, p < .001). The $15,000-$24,999 income bracket was the sole income category that did not differ significantly from the lowest bracket (aOR 0.96, p = .609), suggesting negligible marginal reduction in food insecurity risk between the two lowest income groups. SNAP receipt was associated with more than twice the odds of food insecurity (aOR 2.22, 95% CI 1.99-2.48, p < .001), consistent with SNAP functioning as a marker of material deprivation that is not fully captured by income alone. Higher educational attainment was associated with progressively lower odds of food insecurity, with college graduates having 55% lower odds than those with less than a high school education (aOR 0.45, 95% CI 0.38-0.52, p < .001). Female sex (aOR 1.23, p < .001), unemployment (aOR 1.39, p < .001), inability to work (aOR 1.21, p = .011), fair or poor general health (aOR 1.45, p < .001), greater physical and mental unhealthy days, and a history of depressive disorder (aOR 1.11, p = .047) were each independently associated with higher odds of food insecurity. Older age was associated with lower odds, with adults 65 and older having 53% lower odds than those aged 18-24 (aOR 0.47, 95% CI 0.38-0.59, p < .001). Marital status and urban/rural status were not significant predictors after full adjustment.

To assess whether the association between social support and food insecurity varied by race/ethnicity, fully adjusted models were estimated separately within each racial group, with race excluded from the model formula. Results are presented in Table 4. The association between inadequate social support and food insecurity was significant and consistent across White, Black, and Hispanic respondents, though the magnitude differed across groups. Among White, non-Hispanic respondents, receiving support sometimes or rarely/never was associated with 84% (aOR 1.84, 95% CI 1.60–2.12) and 113% (aOR 2.13, 95% CI 1.81–2.51) higher odds of food insecurity, respectively. Among Black, non-Hispanic respondents, the sometimes estimate was somewhat larger (aOR 2.16, 95% CI 1.75–2.68), while the rarely/never estimate was comparatively attenuated (aOR 1.76, 95% CI 1.35–2.30), suggesting that for this group the association may peak at intermediate levels of support deprivation rather than increasing monotonically. Among Hispanic respondents, effect sizes were smaller but remained significant for both sometimes (aOR 1.55, 95% CI 1.17–2.04) and rarely/never (aOR 1.41, 95% CI 1.02–1.94), indicating that while the association persists, it is weaker relative to other groups, consistent with the lower overall attenuation observed in the fully adjusted pooled model. No significant associations were observed for Other, non-Hispanic respondents across any support level, though this group had the smallest analytic subsample and results should be interpreted with caution. Among Multiracial respondents, the sometimes and rarely/never contrasts were both significant (aOR 1.92 and 2.13 respectively), though confidence intervals were wide reflecting smaller cell sizes. A formal test of interaction between social support and race/ethnicity was non-significant (F = 1.33, p = .194), indicating that observed differences in effect magnitude across groups should not be interpreted as evidence of statistically meaningful effect modification.

**Table 4.**
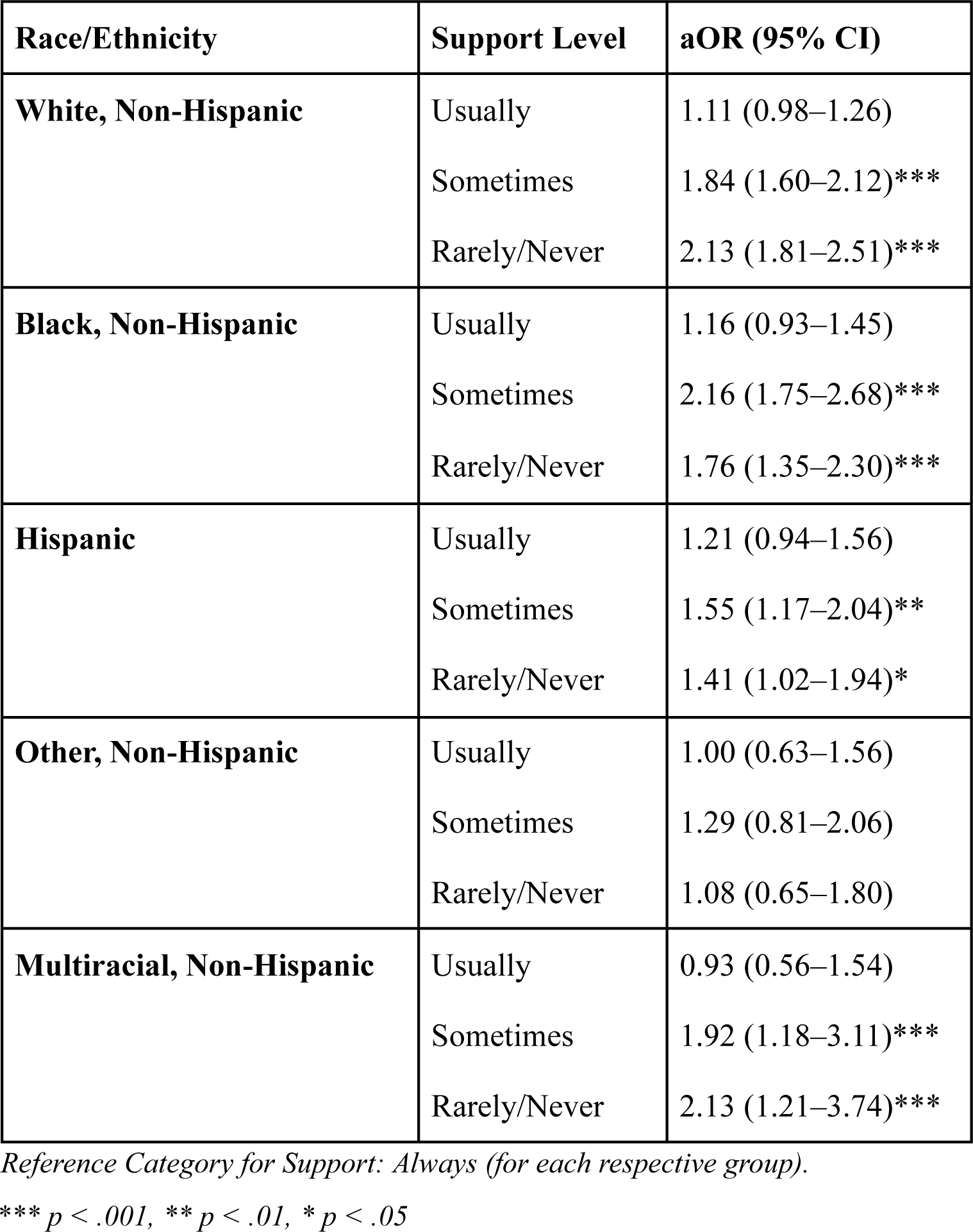
Adjusted ORs by Race/Ethnicity and Support Level.

Urban/rural stratified models were estimated to examine whether the association between social support and food insecurity differed by residential context. Among urban respondents (n = 160,804), receiving support sometimes or rarely/never was associated with 73% (aOR 1.73, 95% CI 1.53–1.95) and 68% (aOR 1.68, 95% CI 1.44–1.95) higher odds of food insecurity, respectively, relative to always receiving support. Among rural respondents (n = 24,472), the corresponding estimates were notably larger – 103% (aOR 2.03, 95% CI 1.58–2.61) and 93% (aOR 1.93, 95% CI 1.46–2.54) higher odd; however, the confidence intervals overlap with the urban results, and thus they should not be considered significantly different (F = 0.42, p = .738). The usually versus always contrast was non-significant in both strata.

## Discussion

This study was, to the best of our knowledge, the first nationwide analysis in the United States to assess the relationship between social and emotional support and food insecurity. Consistent with our hypothesis, inadequate social support was independently associated with significantly elevated odds of food insecurity after controlling for a comprehensive set of sociodemographic, socioeconomic, health, and geographic covariates. The association demonstrated a clear dose-response pattern, with progressively greater odds of food insecurity observed at lower levels of social support, and remained robust across multiple model specifications and sensitivity analyses.

In the fully adjusted primary model, adults who reported sometimes receiving needed social support had 75% higher odds of food insecurity compared to those who always received support, while those who rarely or never received support had 69% higher odds; the confidence intervals overlapped for the sometimes and rarely/never support levels. Notably, the usually versus always contrast was non-significant across all three models, suggesting that the relationship is not strictly linear and that a meaningful threshold exists between adequate and inadequate support rather than across the full ordinal gradient. In the secondary model run with an ordinal food insecurity outcome, the usually versus always contrast became significant (Appendix A). This threshold effect has practical implications, as it suggests that even partial social support may be sufficient to buffer against food insecurity risk.

The racial stratified analyses revealed that the association was significant and consistent among White, Black, and Hispanic respondents, though effect sizes varied meaningfully across groups. The association was largest among White and Multiracial respondents and comparatively attenuated among Hispanic adults, for whom food insecurity may be more heavily determined by structural economic factors than by psychosocial resources. While a formal interaction test did not reach statistical significance, the pattern of variation across groups warrants attention in future research, particularly given evidence that the social determinants of food insecurity operate differently across racial and ethnic communities.^8^

The urban/rural stratified analyses yielded meaningful findings within the sensitivity analyses. The association between inadequate social support and food insecurity was consistently stronger in rural settings, with rural adults who reported sometimes or rarely/never receiving support facing approximately twice the odds of food insecurity compared to those always receiving support (aOR 2.03 and 1.93, respectively), compared to effect sizes of 1.73 and 1.68 among urban adults. As discussed earlier, however, the confidence intervals demonstrated overlap, and the rural/urban difference was not statistically significant. Interestingly, the rural intervals demonstrated higher maximums than the urban intervals. This amplified association in rural areas likely reflects the reduced availability of formal institutional supports – food banks, social services, and community organizations – that characterize many rural communities, rendering informal social networks a more critical resource for food access. These findings are consistent with prior evidence suggesting that social connectedness plays a particularly salient role in buffering food insecurity in rural and resource-limited environments.^6,9^

### Potential Mechanisms

The observed association between inadequate social support and increased food insecurity likely operates through several different interconnected pathways. Drawing from established public health and sociological frameworks, we can propose two primary mechanisms that explain the relationship: the Social Capital Model and the Stress-Buffering Model.

The essence of the social capital model is rooted deeply in the conceptualization of social capital^15^, suggesting that social networks provide resources that can directly and indirectly influence food insecurity.^16^ Social connections serve as a vehicle for both material assistance and information about food resources. For example, individuals with robust social connections may receive either direct food assistance from their families or their friends when they face periods of hardship. Additionally, being a part of a community allows individuals to have a reliable source of information and access resources such as community food pantries or seasonal food distribution events that they might not otherwise discover.

Transportation barriers, which disproportionately affect food-insecure populations^10,11^, may be amended through social networks. Friends, family, and other close social connections could be viable actors to ameliorate transportation barriers to food pantries, grocery stores, or SNAP offices, or even offering shopping arrangements that can ultimately reduce individuals’ transportation costs. This mechanism might be prominent in more rural areas, as geographic isolation causes further food access challenges^12^, and where our stratified analyses suggested a substantial association between social support and food security that is supported by existing qualitative literature.^13^

The informational dimension of the social capital extends beyond knowing about the available resources; it must also encompass the practical, experience-based knowledge individuals must use to navigate food systems equally. For example, the advantage for socially connected individuals could be that they may learn about budget-friendly shopping techniques, bulk purchasing cooperatives, or even community parties. This knowledge transfer is a strong tool that can enhance food purchasing power and diversify food acquisition strategies beyond retail channels.

The Stress-Buffering hypothesis proposes that social support modifies the relationship between economic stressors and their health-related outcomes, as well as food insecurity.^17^ It can be argued that financial strain creates psychological stress that can impair decision-making, reduce motivation, and reduce cognitive ability to maintain successful household management. It has been demonstrated that reminding individuals of their strong social ties increases their perception of food availability^14^; as such, it may also be the case that having positive social connections reduces the psychological burden of food insecurity and results in lower reported/perceived levels thereof.

It is important to note that these mechanisms likely operate bidirectionally. While inadequate social support may increase food insecurity risk, food insecurity itself may erode social connections. The shame and stigma associated with food insecurity may lead individuals to withdraw from social activities, decline social invitations involving food, or avoid situations where their economic circumstances might become apparent.^18^ This social withdrawal can gradually weaken the very networks that could assist, creating a vicious cycle of increasing isolation and food insecurity.

The relative contribution of these mechanisms likely varies across populations and contexts. Our race-stratified analyses revealed differential effect sizes across racial groups, potentially reflecting variations in social network structures, cultural norms around help-seeking^19^, or differential access to formal support systems. Similarly, the slightly stronger associations observed in rural areas may reflect greater reliance on informal social networks in communities with limited formal social services.

### Strengths and Limitations

A significant strength of this study is the use of BRFSS data, which is often credited to be the world’s largest continuously conducted health survey system. Through this dataset, our analysis benefits from a large, nationally representative sample, which significantly enhances the generalizability of our findings across diverse geographic and socioeconomic strata in the United States. Furthermore, the standardized data collection protocols of the BRFSS minimize measurement variance, allowing for a robust assessment of the intersection between social support and food insecurity risk.

Despite these strengths, several limitations must be acknowledged. First, the cross-sectional design of the BRFSS prevents causal inferences between food insecurity and social support to be definitively determined. Second, the data may be subject to non-response bias, particularly within rural populations. Telephone-based surveys often face lower participation rates in rural areas due to factors such as connectivity issues, which leads to an unclear estimation of the challenges faced by the most isolated communities. Similarly, the geographic granularity of this study was limited by the use of the NCHS Six-Level Urban-Rural Classification scheme. While the NCHS 6-tier definition is a validated standard for health research, it is less expansive than other metrics (such as the USDA’s Rural-Urban Commuting Area codes). Consequently, our model may not capture the full nuance of rurality, potentially masking disparities during analysis.

### Future Directions

The findings presented here raise several questions that warrant further investigation, both to clarify the mechanisms underlying the observed associations and to translate these findings into actionable interventions.

The cross-sectional design of the BRFSS precludes causal inference, and future work should prioritize longitudinal approaches capable of establishing temporal precedence between social support erosion and food insecurity onset. Cohort studies tracking changes in social connectedness alongside food security status over time would allow researchers to distinguish whether inadequate social support precedes food insecurity, follows from it, or operates bidirectionally, as stress-process models would suggest. There is also a need for qualitative inquiry; while the regression estimates reported here are consistent with the hypothesis that social support operates as a protective buffer against food insecurity, they cannot confirm the specific pathways through which this association operates. Interviews and focus group research with food-insecure individuals, particularly those within communities with varying levels of social infrastructure, would help identify whether the primary mechanisms are informational (e.g., awareness of food assistance programs), instrumental (e.g., resource sharing and mutual aid), or emotional (e.g., stress buffering that preserves decision-making capacity). Confirming these mechanisms is needed before designing interventions that target the right leverage points.

Beyond mechanism identification, future research should attempt to quantify the relative contribution of distinct social support pathways to the overall association. Formal mediation analyses incorporating measures of perceived stress, network density, and access to tangible aid would allow researchers to decompose the total effect of social support on food insecurity into its constituent indirect pathways. This analysis would be particularly valuable for racial and rural subgroups, where the magnitude of the association differed across this study’s stratified models, suggesting that the mechanisms themselves may not be uniform across populations.

Finally, the policy and programmatic implications of this work deserve direct empirical attention. Existing food assistance programs, including SNAP, are designed primarily around income eligibility and do not formally integrate social support as either a screening criterion or a program component. Intervention studies evaluating whether co-locating food assistance with social support services, peer navigation programs, or community health worker models meaningfully reduces food insecurity would substantially advance this literature. Evaluation of such programs using quasi-experimental designs would generate the kind of causal evidence needed to inform resource allocation at the state and federal level, particularly in rural communities where the buffering role of social connectedness appears to be most pronounced.

## Acknowledgements

We wish to thank Dr. Ryan Talbert, Department of Sociology, University of Connecticut, for his guidance and support in carrying out this project.

## CRediT

Eashwar Krishna *(Conceptualization, Data Curation, Formal Analysis, Investigation, Methodology, Supervision, Validation, Writing - original draft, Writing - review & editing)*, Neha Shanavas *(Conceptualization, Visualization, Writing - original draft, Writing - review & editing)*, Fatima Mir (*Project Administration, Methodology, Writing - original draft, Writing - review & editing)*, Ahan Kothapeta (*Conceptualization, Writing - original draft, Writing - review & editing)*, Carlos Duluc (*Writing - original draft, Writing - review & editing)*, Roma Kale *(Writing - original draft, Writing - review & editing)*, Priya Bheemanakunta *(Writing - original draft, Writing - review & editing)*, Eshita Mathur *(Conceptualization, Writing - original draft, Writing - review & editing)*

## Data Availability

The script can be found in this repository and the data at the CDC BRFSS website.

## Appendix A

**Table A1.).**

| Support Level | aOR (95% CI) | p-value |
| --- | --- | --- |
| Usually | 1.27 (1.17–1.38) | <0.001 |
| Sometimes | 1.73 (1.57–1.89) | <0.001 |
| Rarely/Never | 1.70 (1.51–1.91) | <0.001 |

*Reference Category for Support: “Always”. Food insecurity was coded as 4 ordinal tiers (always, sometimes, usually, or rarely/never having food bought not last and not having money to buy more). Odds ratios represent chances of being in a tier one stage up.*

